# Assessing clinical outcomes of vancomycin treatment in adult patients with penicillin-resistant *Enterococcus faecium* bacteremia

**DOI:** 10.1101/2022.04.27.22274384

**Authors:** Hiroshi Sasano, Ryutarou Arakawa, Kazuhiko Hanada

**Author notes:** Corresponding author (KH).

## Abstract

Enterococcal bacteremia is associated with high mortality and long-term hospitalization. Here, we aimed to investigate the clinical outcomes and evaluate risk factors for mortality in adult patients treated with vancomycin (VCM) for penicillin-resistant *Enterococcus faecium* (*E. faecium*) bacteremia. Data were collected from inpatients at a single university hospital between January 2009 and December 2020. The area under the curve (AUC) of VCM was calculated using the Bayesian approach. The primary outcome was 30-day in-hospital mortality. Univariate analysis showed significant differences in the combined use of vasopressors, history of the use of inactive antimicrobial agents against *E. faecium*, VCM plasma trough concentration, and renal dysfunction during VCM administration between 30-day mortality and survival groups. However, the AUC/minimum inhibitory concentration (MIC) was not significantly different. Multivariate analysis revealed that concomitant vasopressors were an independent risk factor for 30-day all-cause mortality (odds risk, 7.81; 95% confidence interval, 1.16–52.9; *P* = 0.035). VCM plasma trough concentrations and AUC/MIC in the mortality group were higher than those in the surviving group. Depending on the drug-susceptibility results of *E. faecium*, it is considered that the AUC/MIC is calculated at a double AUC value. No association between AUC/MIC and treatment effect in *E. faecium* bacteremia was assumed because the known target AUC/MIC was sufficiently achieved in the mortality group. When an immunocompromised host develops *E. faecium* bacteremia with septic shock, especially in situations where the hemodynamics of using a pressor agent is unstable, treating it exclusively by sufficient exposure to antibacterial agents may be difficult.

## Introduction

*Enterococcus* species are gram-positive facultative anaerobic cocci that constitute the normal bacterial flora in human and animal intestines. However, enterococcal bacteremia is associated with high mortality and long-term hospitalization [1-3]. Most cases of invasive enterococcal bacteremia are caused by *Enterococcus faecalis*, followed by *Enterococcus faecium* (*E. faecium*) [4]. *E. faecium* is commonly found in the microflora of the human gut and mainly causes urinary tract infections, wound infections, endocarditis, and bacteremia [5].

Vancomycin (VCM) is used as a first-line drug for the treatment of **penicillin-resistant** *E. faecium* infections to achieve clinical efficacy and maintain minimum toxicity, and the area under the serum concentration-time curve (AUC)/minimum inhibitory concentration (MIC) ratio of VCM is recommended to be 400–600. However, this target ratio is considered for the suspected or definitive diagnosis of methicillin-resistant *Staphylococcus aureus* infections [6]. A previous study suggested that an AUC/MIC ≥389 was an independent factor in reducing mortality from enterococcal bacteremia [7]. However, that study evaluated all *Enterococcus* spp. and was not limited to *E. faecium*, and evaluation was performed by VCM treatment for bacterial species, such as *E. faecium*, that could be treated with penicillin antimicrobial agents. To date, the effects of disease status, concomitant agents, and pharmacokinetic/pharmacodynamic parameters, such as plasma trough concentration and AUC/MIC, on the clinical outcomes of VCM administration have not yet been investigated. Thus, in this study, we aimed to investigate the clinical outcomes and evaluate risk factors for mortality in adult patients treated with VCM for penicillin-resistant *E. faecium* bacteremia.

## Materials and methods

### Study population

Data were collected from inpatients of Juntendo University Hospital between January 2009 and December 2020. The study was conducted in accordance with the guidelines of the Helsinki declaration, and the protocol was approved by the ethics committee of our hospital (approval no. 20-270). To treat the existing information in the medical records retrospectively, instead of omitting informed consent, information on the implementation of this study was posted on the homepage of the Juntendo University Hospital Department of Pharmacy for the patients and their families to be studied, thus providing them with the opportunity to refuse to participate.

The inclusion criteria were as follows: detection of *E. faecium* in blood culture and catheter tip culture of the patient, administration of VCM to treat bacteremia, plasma VCM concentration data, and age <18 years. The exclusion criteria were as follows: administration of renal replacement therapy (hemodialysis or continuous hemodiafiltration), no plasma VCM concentration data, lack of laboratory data, administration of antimicrobial agents other than VCM for the treatment of *E. faecium* bacteremia, isolation of blood cultures with the simultaneous detection of gram-positive bacterial species other than *E. faecium*, isolation of blood cultures with VCM-resistant *E. faecium* (VRE), penicillin-resistant *E. faecium, E. faecium* not treated as a contaminating bacterium, and no MIC data.

Only the first episode of *E. faecium* bacteremia was considered. Patients with polymicrobial bacteremia were only included in the analysis if they had received antimicrobial agents that were active in vitro against other coinfection pathogens.

The following patient data were extracted from medical records using predesigned forms: demographic characteristics (body weight, height, age, sex, underlying diseases, and admission to the intensive care unit), diagnosis of infectious diseases and source of infection, clinical laboratory data (levels of alanine aminotransferase, alkaline phosphatase, serum creatinine, and total bilirubin; and platelet and white blood cell counts), 30-day all-cause mortality, dose of VCM administered, interval and initial plasma trough concentration of VCM at steady state, concomitant agents, and MIC of *E. faecium*. The onset of bacteremia was defined as the date of collection of the first blood culture that was positive for *E. faecium*. Regarding the drugs taken before the onset of *E. faecium* bacteremia, the following cases were defined as “use”: steroids and antineoplastic agents within 30 days before the positive blood culture; antibacterial drugs, when administered continuously for at least 48 h/month before positive blood culture.

### Calculation of AUC

The AUC from 0 to 24 h (AUC_24_) of VCM was calculated using the Bayesian approach performed with therapeutic drug monitoring software (Vancomycin MEEK TDM analysis software ver 3.0; Meiji Seika Pharma Co., Ltd., Tokyo, Japan). Thereafter, the AUC_24_/MIC ratio was calculated using the AUC_24_ obtained for each patient and the MIC derived from the microbiological test results.

### Measurement of MIC of E. faecium

The MIC was measured using an automated microdilution method.

### Primary and secondary endpoints

The primary outcome measure was 30-day in-hospital mortality [7]. The secondary endpoints were a microbiological failure, defined as the detection of the same bacterial species using blood culture retest during treatment with VCM, and recurrence of *E. faecium* bacteremia within 90 days after treatment of bloodstream infections due to *E. faecium* from negative blood culture. Such instances were defined as complex re-positive cultures. The presence or absence of other factors in relation to renal dysfunction was also examined, and renal disorders were evaluated using the International Clinical Practice Guidelines [8]. These endpoints were compared between the two groups with and without recurrence of *E. faecium* bacteremia.

### Statistical analyses

Univariate data were analyzed using the Mann–Whitney U test for continuous variables and Fisher’s exact test for categorical variables. In the multivariate analysis, the correlation between the two variables was analyzed using Spearman’s ranking method. Among the items for which no correlation was found, those deemed to be related to the patient’s condition leading to death and the concomitant agents associated with the onset of renal dysfunction during VCM administration were analyzed using logistic regression analysis. Statistical significance was set at *P* <0.05.

## Results

Of the 64 patients diagnosed with *E. faecium* bacteremia during the study period, 41 were enrolled in this study **(Table 1)**. The median age was 69 years, and the creatinine clearance calculated using the Cockcroft–Gault formula was 59.4 mL/min. Overall, 51.2% (21/41) of patients had a history of admission to the intensive care unit, and 63.4% (26/41) of patients had a malignant tumor as the underlying disease. Biliary tract infection was the most common disease (36.6%) among the patients. The VCM MIC value was the highest at concentrations ≤0.5 μg/mL (20 strains), followed by that at concentrations of 1 μg/mL (16 strains). Nine of the 41 patients (22.0%) died within 30 days of diagnosis with *E. faecium* bacteremia and were classified in the “30-day in-hospital mortality group.” Meanwhile, 32 patients were classified in the “survival group.”

**Table 1.**
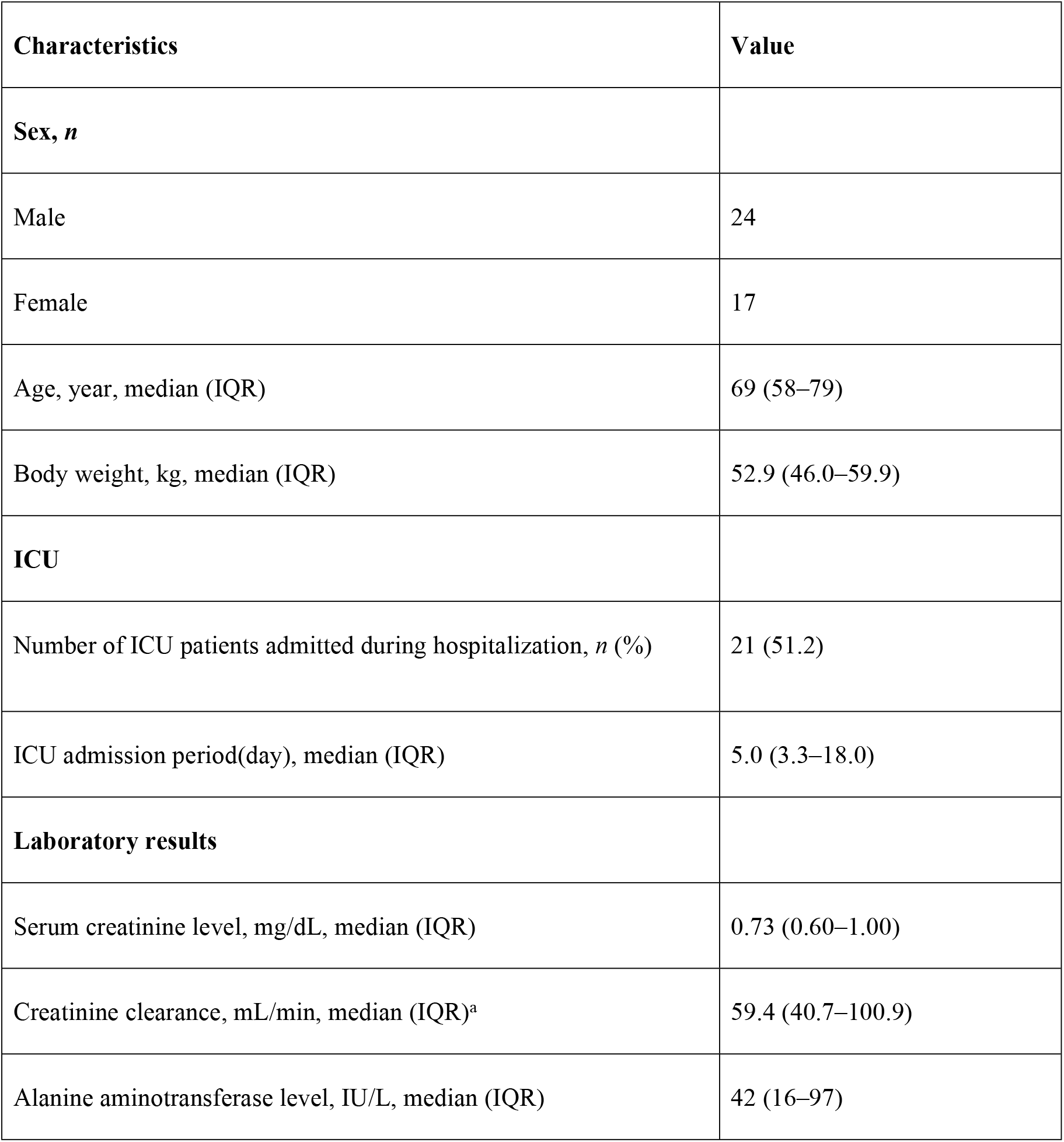
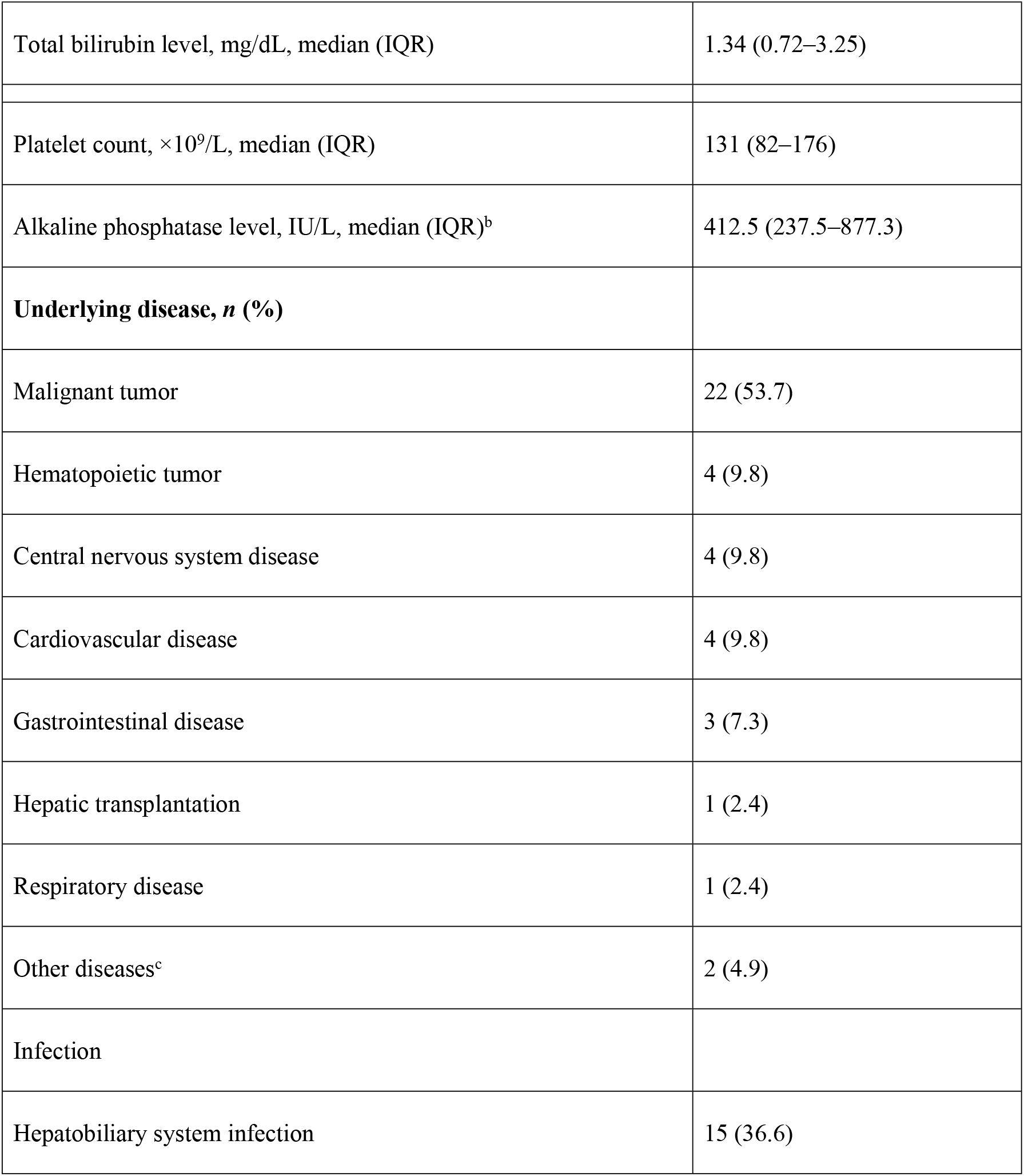
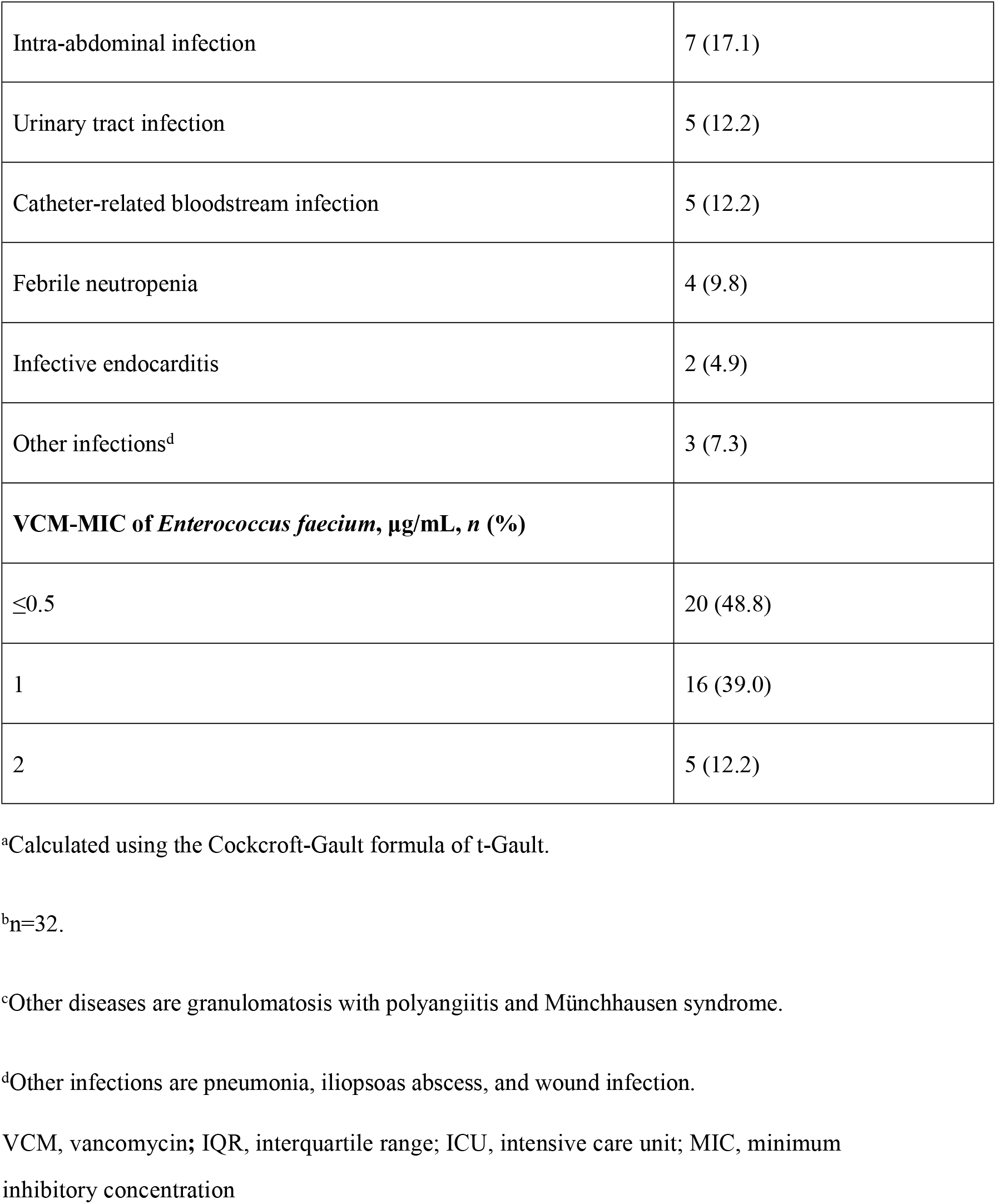
Patient characteristics.

| **Characteristics** | **Value** |
| --- | --- |
| **Sex, *n*** |  |
| Male | 24 |
| Female | 17 |
| Age, year, median (IQR) | 69 (58–79) |
| Body weight, kg, median (IQR) | 52.9 (46.0–59.9) |
| **ICU** |  |
| Number of ICU patients admitted during hospitalization, *n* (%) | 21 (51.2) |
| ICU admission period(day), median (IQR) | 5.0 (3.3–18.0) |
| **Laboratory results** |  |
| Serum creatinine level, mg/dL, median (IQR) | 0.73 (0.60–1.00) |
| Creatinine clearance, mL/min, median (IQR)<sup>a</sup> | 59.4 (40.7–100.9) |
| Alanine aminotransferase level, IU/L, median (IQR) | 42 (16–97) |
| Total bilirubin level, mg/dL, median (IQR) | 1.34 (0.72–3.25) |
| Platelet count, $\times 10^9/\text{L}$, median (IQR) | 131 (82–176) |
| Alkaline phosphatase level, IU/L, median (IQR)<sup>b</sup> | 412.5 (237.5–877.3) |
| **Underlying disease, *n* (%)** |  |
| Malignant tumor | 22 (53.7) |
| Hematopoietic tumor | 4 (9.8) |
| Central nervous system disease | 4 (9.8) |
| Cardiovascular disease | 4 (9.8) |
| Gastrointestinal disease | 3 (7.3) |
| Hepatic transplantation | 1 (2.4) |
| Respiratory disease | 1 (2.4) |
| Other diseases<sup>c</sup> | 2 (4.9) |
| Infection |  |
| Hepatobiliary system infection | 15 (36.6) |
| Intra-abdominal infection | 7 (17.1) |
| Urinary tract infection | 5 (12.2) |
| Catheter-related bloodstream infection | 5 (12.2) |
| Febrile neutropenia | 4 (9.8) |
| Infective endocarditis | 2 (4.9) |
| Other infections<sup>d</sup> | 3 (7.3) |
| **VCM-MIC of *Enterococcus faecium*, µg/mL, n (%)** |  |
| ≤0.5 | 20 (48.8) |
| 1 | 16 (39.0) |
| 2 | 5 (12.2) |

<sup>a</sup>Calculated using the Cockcroft-Gault formula of t-Gault.
<sup>b</sup>n=32.
<sup>c</sup>Other diseases are granulomatosis with polyangiitis and Münchhausen syndrome.
<sup>d</sup>Other infections are pneumonia, iliopsoas abscess, and wound infection.
VCM, vancomycin; IQR, interquartile range; ICU, intensive care unit; MIC, minimum
inhibitory concentration

Univariate analysis showed significant differences in the concomitant use of vasopressors, use of inactive antimicrobial agents against *E. faecium*, VCM plasma trough concentration, and renal dysfunction between 30-day in-hospital mortality and survival groups during VCM administration (**Table 2**). However, the AUC_24_/MIC values in the two groups were not significantly different. We then performed a multivariate analysis on two uncorrelated variables, namely renal dysfunction during VCM administration and the combined use of vasopressors. We identified the combined use of vasopressors as an independent risk factor for 30-day all-cause mortality (**Table 3**).

**Table 2.** Comparison between 30-day mortality and survival groups.

| Variable | Survival (n=32) | 30-day mortality<br>(n=9) | *P* |
| --- | --- | --- | --- |
| **Disease status, *n* (%)** |  |  |  |
| Solid tumor | 18 (56.3) | 4 (44.4) | 0.71 <sup>a</sup> |
| Digestive system tumor | 17 (53.1) | 3 (33.3) | 0.45 <sup>a</sup> |
| Hepatobiliary tumor | 11 (34.4) | 2 (22.2) | 0.69 <sup>a</sup> |
| Diabetes mellitus | 5 (15.6) | 3 (33.3) | 0.34 <sup>a</sup> |
| ICU admission | 15 (46.9) | 6 (66.7) | 0.45 <sup>a</sup> |
| Use of central intravenous catheter | 12 (37.5) | 6 (66.7) | 0.15 <sup>a</sup> |
| Hepatobiliary system infection | 12 (37.5) | 3 (33.3) | 1.00 <sup>a</sup> |
| Intra-abdominal infection | 7 (21.9) | 0 (0.0) | 0.31 <sup>a</sup> |
| Urinary-tract infection | 3 (9.4) | 2 (22.2) | 0.30 <sup>a</sup> |
| Catheter-related bloodstream infection | 2 (6.25) | 3 (33.3) | 0.06 <sup>a</sup> |
| Infective endocarditis | 2 (6.3) | 0 (0.0) | 1.00 <sup>a</sup> |
| **Use of concomitant agents, *n* (%)** |  |  |  |
| Thrombomodulin | 3 (9.4) | 0 (0.0) | 1.00 <sup>a</sup> |
| Vasopressor | 3 (9.4) | 5 (55.6) | 0.007<br><sup>a</sup> |
| Globulin | 2 (6.3) | 2 (22.2) | 0.20 <sup>a</sup> |
| Total parenteral nutrition | 14 (43.8) | 3 (33.3) | 0.71 <sup>a</sup> |
| **Medication history, *n* (%)** |  |  |  |
| Steroids<sup>c</sup> | 9 (28.1) | 4 (44.4) | 0.43 <sup>a</sup> |
| Anticancer agents<sup>c</sup> | 7 (21.9) | 2 (22.2) | 1.00 <sup>a</sup> |
| Antimicrobial agents that are inactive against *E. Faecium*, including cephalosporins and carbapenems<sup>d</sup> | 13 (40.6) | 8 (88.9) | 0.02 <sup>a</sup> |
| Antimicrobial agents<sup>d</sup> | 15 (46.9) | 8 (88.9) | 0.05 <sup>a</sup> |
| **VCM administration, median (IQR)** |  |  |  |
| Duration, day | 15 (13–20.3) | 7 (6–16) | 0.05 <sup>b</sup> |
| Dose, mg/day | 1250 (1000–2000) | 1000 (750–100) | 0.16 <sup>b</sup> |
| **Pharmacokinetic/pharmacodynamic parameters, median (IQR)** |  |  |  |
| Plasma trough VCM concentration, mg/L | 9.1 (6.6–11.9) | 14.9 (11.0–19.5) | 0.01 <sup>b</sup> |
| AUC/MIC | 658.5 (427.1– | 852 (481.8– | 0.28 <sup>b</sup> |
|  | 990.5) | 1558.0) |  |
| MIC | 1.0 (0.5–1.0) | 0.5 (0.5–1.0) | 0.96 <sup>b</sup> |
| Renal dysfunction during VCM<br>administration | 6 (18.8) | 6 (66.7) | 0.01 <sup>a</sup> |

<sup>a</sup>Fisher's exact test.
<sup>b</sup>Mann–Whitney U test.
<sup>c</sup>Number of uses within 30 days before a positive blood culture test.
<sup>d</sup>Continuous administration for $\geq 48$ hours a month before a positive blood culture.
AUC, area under the curve; VCM, vancomycin; IQR, interquartile range; ICU, intensive care
unit

**Table 3.**
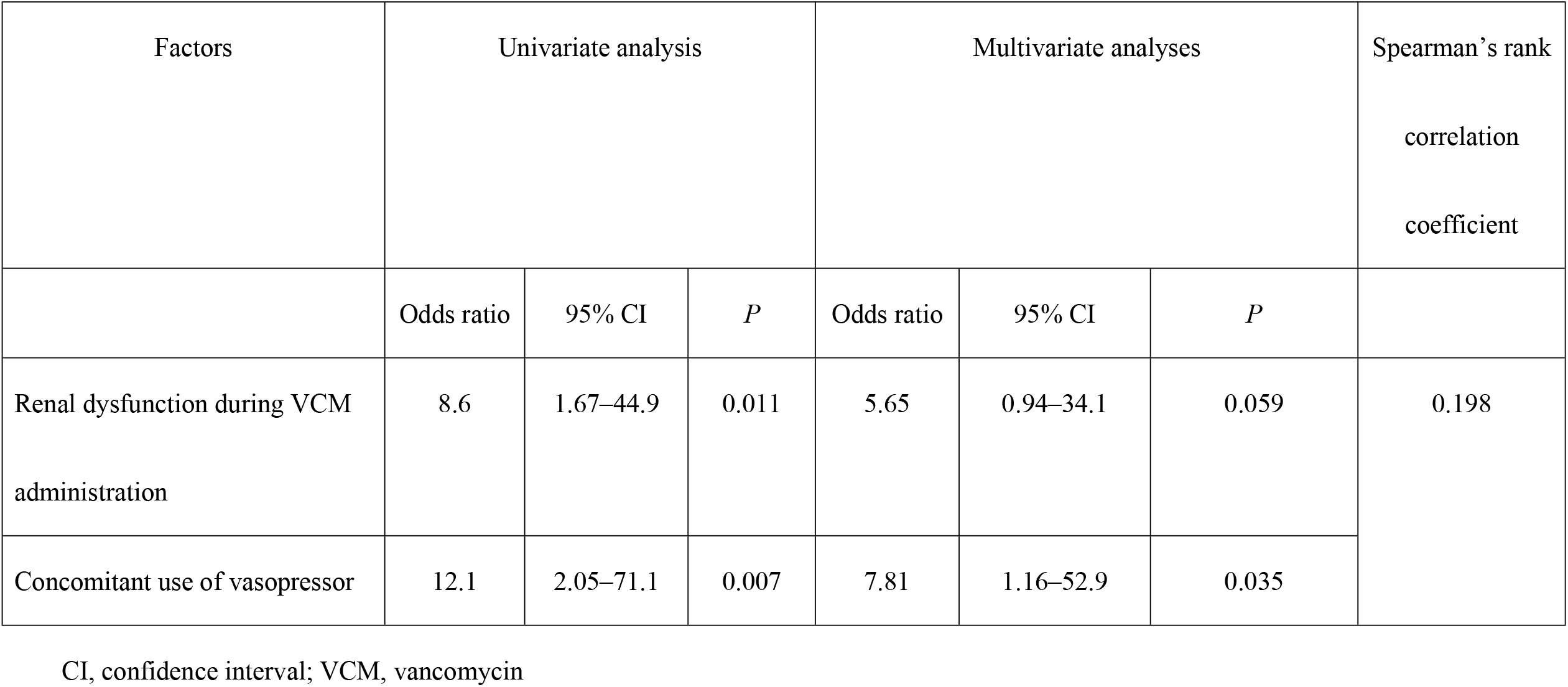
Univariate and multivariate analyses of factors affecting 30-day mortality.

| Factors | Univariate analysis |  |  | Multivariate analyses |  |  | Spearman's rank<br>correlation<br>coefficient |
| --- | --- | --- | --- | --- | --- | --- | --- |
|  | Odds ratio | 95% CI | *P* | Odds ratio | 95% CI | *P* |  |
| Renal dysfunction during VCM<br>administration | 8.6 | 1.67–44.9 | 0.011 | 5.65 | 0.94–34.1 | 0.059 | 0.198 |
| Concomitant use of vasopressor | 12.1 | 2.05–71.1 | 0.007 | 7.81 | 1.16–52.9 | 0.035 |  |

CI, confidence interval; VCM, vancomycin

A comparison of the factors associated with microbiological failures revealed that only the combination of thrombomodulin with VCM had significant results (**see Supplementary Table S1**). The group with renal dysfunction had significantly higher concomitant use of vasopressor and piperacillin-tazobactam (PIPC/TAZ) than without renal dysfunction (**see Supplementary Table S2**) according to the univariate analysis comparison of the presence or absence of renal dysfunction during VCM administration. We performed a multivariate analysis of the factors related to renal dysfunction during VCM administration (vasopressor, PIPC/TAZ, and diuretics) and found that the combination of PIPC/TAZ with VCM was an independent risk factor for renal dysfunction (**see Supplementary Table S3**).

## Discussion

The reported mortality in patients with *E. faecium* bacteremia was 25.0–34.6%, which was similar to the 30-day in-hospital mortality rate (22.0%) observed in this study [9, 10]. In a retrospective study by Jumah et al., the plasma VCM concentrations in enterococcal bacteremia patients were lower than those recommended for *S. aureus* bacteremia, and the mortality groups had lower plasma concentration levels than the survival groups [7]. They also reported that an AUC24/MIC ratio >389 of VCM might reduce mortality due to enterococcal bacteremia [7]. However, in our study, the plasma trough concentrations and the AUC24/MIC ratio of VCM in the mortality group were higher than those in the surviving group, suggesting that the known plasma trough concentrations and AUC24/MIC ratio may not be associated with mortality in *E. faecium* bacteremia patients. We speculate that when the drug-susceptibility of *E. faecium* is determined to be 0.5 μg/mL, the AUC/MIC is calculated as a double value of the AUC. The MIC of *E. faecium* in 5 patients (56%) in the mortality group was 0.5 μg/mL. Among them, one had an AUC of 426 mg per h/L and an AUC/MIC ratio of 852 according to the Bayesian method, whereas the other 4 patients had an AUC value >400 mg per h/L and an AUC/MIC ratio of >800. Thus, the known target AUC/MIC was sufficiently achieved even in the mortality group. These results suggest that the AUC/MIC ratio is not associated with *E. faecium*.

Another possibility is that VCM is bactericidal against most gram-positive bacteria but bacteriostatic against enterococci. [11]. Bacteriostatic antibacterial agents require phagocytic cells to eliminate bacteria and may be less effective in patients without a normal immune response [12]. Therefore, the known AUC24/MIC ratio >400 may not be a therapeutic target for enterococci. In univariate analysis, plasma VCM concentration, renal dysfunction during VCM administration, use of other antimicrobial agents inactive against *E. faecium* (such as cephalosporins), and the concomitant use of vasopressors were identified as factors affecting mortality. In multivariate analysis, the concomitant use of vasopressors emerged as an independent factor affecting mortality. When the host’s body exhibits a decrease in immune function due to a malignant tumor or the like, sepsis tends to become severe and causes shock. The International Guidelines for the Management of Septic Shock 2016 recommend the use of vasopressors for a septic shock when such patients do not respond to initial fluid administration [13]. Moreover, a study predicting the risk of VCM-induced acute kidney injury (AKI) reported that 32 (36.8%) of 87 patients who received vasopressors developed AKI and were being treated [14]. The development of AKI is thought to increase the risk of death from an enterococcal bloodstream invasion [15].

According to medical information records, 11 of 41 target patients were diagnosed with septic shock, and eight were concomitantly treated with vasopressors. In the group of patients who died within 30 days, four of five patients who received a pressor agent developed renal dysfunction. Previous studies investigating the predictors of *S. aureus*-related mortality within 30 days have similarly reported that septic shock increases mortality, which is consistent with our results [16, 17]. However, the concomitant use of vasopressors as a risk factor for *E. faecium* bacteremia is a novel finding of this study. A higher prevalence of sepsis has been reported in oncological patients vs. non-oncological patients [18]. Immunosuppression due to underlying malignancy or its treatment can increase the risk for severe infections [19]. Sixty-five percent of the patients included in this study had a background of immunosuppressive conditions such as solid tumors, hematological tumors, and liver transplantation, and all patients in the 30-day mortality group had a background of malignant tumors. Therefore, when an immunocompromised host develops *E. faecium* bacteremia with septic shock, especially in situations where the hemodynamics of using a pressor agent is unstable, it may be difficult to treat exclusively by sufficient exposure to antibacterial agents.

Moreover, univariate analysis showed that using thrombomodulin in combination with VCM was a risk factor for microbiological failure at the secondary endpoint. As a type of conventional biological defense mechanism, thrombus formation contributes to resistance to bacterial infections. However, sepsis can cause disseminated intravascular coagulation owing to an unregulated state of the biological defense mechanism. In other words, in patients where thrombomodulin therapy is used in addition to antimicrobial agents, treatment of bloodstream infections may become challenging because disseminated intravascular coagulation presents as the formation of excessive thrombi throughout the body. Furthermore, the combined use of PIPC/TAZ and VCM was identified as a risk factor for renal dysfunction. Although the increased risk of renal disorder due to the concomitant use of PIPC/TAZ and VCM has been previously reported, the results are consistent [20, 21].

This study has several limitations that are inherent to its retrospective design. The number of cases was very limited due to the single university hospital and the target of penicillin-resistant *E. faecium* bacteremia cases.

In addition, in the 30-day mortality group, the high AUC/MIC may indicate that the systemic exposure increased because of the delayed excretion due to the decreased renal function. However, it could not be defined accurately owing to various factors such as concomitant agents, decreased perfusion to the body’s organs associated with sepsis or septic shock, and administration of vancomycin. For these reasons, we cannot exclude the possibility that residual bias affected our findings.

In conclusion, immunosuppressive patients with penicillin-resistant *E. faecium* bloodstream infections undergoing treatment with vasopressors with unstable hemodynamics due to septic shock may be difficult to treat with sufficient antimicrobial exposure alone. However, VCM was the most effective drug for penicillin-resistant *E. faecium*. Therefore, it is necessary to consider concomitant drugs and provide suitable treatment individually for each patient while monitoring blood concentration to avoid the onset of renal dysfunction.

## Data Availability

All relevant data are within the manuscript and its Supporting Information files.

## Statements and Declarations

## Author Contributions

All authors contributed to the study’s conception and design. Material preparation, data collection, and analyses were performed by HS. The first draft of the manuscript was written by HS, and all authors commented on previous versions of the manuscript. All authors read and approved the final manuscript.

## Data Availability

The datasets generated and analyzed during the current study are available from the corresponding author upon reasonable request.

## Supporting Information

**S1 Table. Comparison between groups with and without microbiological failure**

^a^Fisher’s exact test.

^b^Mann–Whitney U test.

^c^Number of uses within 30 days before a positive blood culture test.

^d^Continuous administration for ≥48 hours a month before a positive blood culture.

AUC, area under the curve; VCM, vancomycin; IQR, interquartile range; MIC, minimum inhibitory concentration

**S2 Table. Comparison between groups with and without renal dysfunction during VCM administration**.

^a^Fisher’s exact test.

^b^Mann–Whitney U test.

^c^Number of uses within 30 days before a positive blood culture test.

^d^Continuous administration for ≥48 hours a month before a positive blood culture.

AUC, area under the curve; VCM, vancomycin; IQR, interquartile range; PIPC/TAZ, piperacillin-tazobactam

**S3 Table. Univariate and multivariate analyses of factors affecting renal dysfunction in patients administered VCM**.

CI, confidence interval; PIPC/TAZ, piperacillin-tazobactam; VCM, vancomycin

